# Novel Deep Learning Methodology for Automated Classification of Adamantinomatous Craniopharyngioma Using a Small Radiographic Dataset

**DOI:** 10.1101/2020.04.16.20063057

**Authors:** Eric Prince, Ros Whelan, David M. Mirsky, Todd C. Hankinson

## Abstract

Modern Deep Learning (DL) networks routinely achieve classification accuracy superior to human experts, leveraging scenarios with vast amounts of training data. Community focus has now seen a shift towards the design of accurate classifiers for scenarios with limited training data. Such an example is the uncommon pediatric brain tumor, Adamantinomatous Craniopharyngioma (ACP). Recent work has demonstrated the efficacy of Transfer Learning (TL) and novel loss functions for the training of DL networks in limited data scenarios. This work describes a DL approach utilizing TL and a state-of-the-art custom loss function for predicting ACP diagnosis from radiographic data, achieving performance (CT AUPR=0.99±0.01, MRI AUPR=0.99±0.02) superior to reported human performance (0.87).

## Main

Deep Learning (DL) is a popular machine learning subtype, which is capable of handling advanced classification tasks. Major annual competitions, such as the ImageNet Large Scale Visual Recognition Challenge (ILSVRC),^1^ help advance premier DL models for immensely large datasets. Within the healthcare space, reliable DL inference models have been described under conditions when vast amounts of training data are available. Examples include dermatological diseases and diabetic retinopathy.^1-3^

However, when DL models are trained on more limited datasets, the results are often unreliable, as the models overfit the training data. This is because, in a small-data context, the latent features that a network models likely result from sampling noise that exists in the training data, but not in novel test data^3^. Without techniques to overcome this generalization problem, DL models would be unreliable for less common diseases, including brain tumors, which have limited volumes of data available for model training. As such, the obvious potential for DL models to improve both clinical care and research for rare diseases merits the development of methods to overcome challenges associated with generalization.

One, now canonical, method to overcome the challenge of overfitting small training datasets is Transfer Learning (TL). This is a machine learning methodology that stores knowledge gained from solving a problem within one domain and applies that knowledge to another domain.^4,5^ The success of TL has led to the development of publicly available pre-trained models derived from top DL competition (e.g., ILSVRC) solutions. By using these pre-trained networks to generate informationally rich feature embeddings applicable to our (small-scale) dataset of interest, we enable our classifier to leverage the pattern recognition capabilities of these state-of-the-art architectures.

Another interesting approach for handling small-scale DL classification is to modify the objective function being trained, otherwise known as the loss function. In large-scale contexts, the classic choice for loss function is known as categorical cross-entropy, which is an information model based on the physical principle. Effectively, in this scenario, the DL model is optimized to minimize the distributional distance between predictions and ground-truth labels. This approach is appropriate in large-scale contexts due to the fact that randomly sampling a global distribution repetitively will eventually approximate the true global distribution (i.e., lim_*n*→*N*_ *F*_*X,local*_(*x*_*n*_) = *F*_*X,global*_ (*x*)). In the small-scale context, however, repeat random sampling may never yield a valid approximation of the true global distribution because a random over- or underrepresentation of a given feature within a small dataset is less likely to be overcome through the sheer volume of samples.

To address this, multiple groups have developed novel loss functions designed to yield more generalized discriminative features that are insensitive to outliers.^6-9^ This work investigates the effectiveness of a three-term loss function comprised of (1) sigmoid focal cross-entropy, which weights towards poorly classified training examples and ignores well-classified examples;^8^ (2) triplet hard-loss, which encourages inter-class feature embedding clusters to have minimized distances and intra-class clusters to have maximized distances;^7^ and (3) CORellation ALignment (CORAL) which penalizes for distances between source and target data.^9^ We hypothesize that the concatenation of these terms, along with optimizable trade-off parameters, will afford a loss function that resists overfitting and facilitates increased model capacity (i.e., generalizability).

An additional challenge in identifying the optimal model is the optimization of DL hyperparameters. This remains a complicated and computationally intensive task.^5^ To mitigate the computational time required, one may apply a meta-heuristic parameter optimization in the form of an asynchronously parallelized genetic algorithm. This optimization procedure allows the model to optimize more intelligently over the solution space with fewer required iterations.

Herein, we present a novel Long Short-Term Memory (LSTM) classification framework that achieves radiologist-level classification of non-contrast Computed Tomography (CT) and contrast-enhanced T1-weighted Magnetic Resonance (MR) imaging of ACP against other similar sellar/suprasellar masses that would be considered in the standard radiographic differential diagnosis. Additionally, we demonstrate the importance of the three-loss terms in mitigating the bias-variance tradeoff that is amplified in limited-data instances. This work contributes a transferable and scalable computational framework for statistical inference of rare diseases. Clinical translation will be easily achieved and will improve patient care by reducing the likelihood of misdiagnosis and by allowing for more informed initial diagnostic interactions between patients and clinicians. Furthermore, stepwise advances will lead to a reduction in the need for invasive neurosurgical tissue diagnosis in some clinical circumstances, thereby sparing patients invasive operations on the brain.

## Results

### Network Optimization and Performance

Following parameter optimization via simple Genetic Algorithm (GA), sequence-based classifier for CT and MRI yielded a max AUPR of 0.991 and 0.988, respectively. Additionally, we observed at least 5 network solutions reaching greater than 0.97 AUPR in each modality (Figure 1a-b). Norris, et al. previously demonstrated human experts in a similar scenario achieved a prediction accuracy of 0.87. ^10^

**Figure 1.**
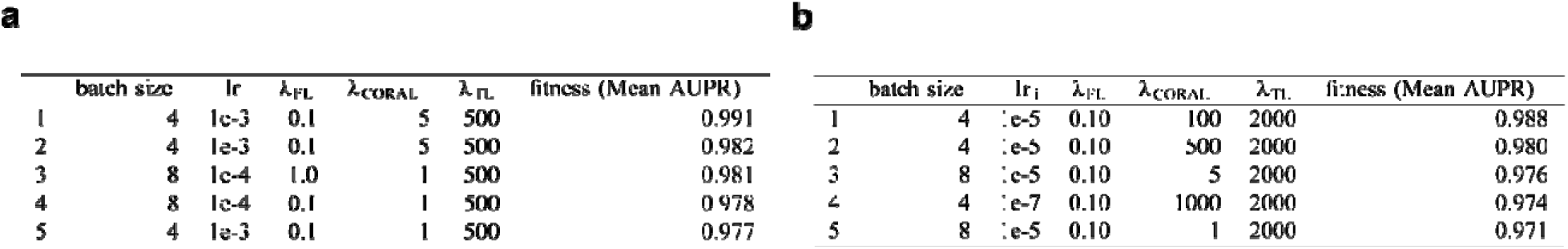
Genetic Algorithm Optimization of Classifier. **a**. Top-5 fittest (highest mean AUPR) CT classifier parameter solutions presented in descending fitness order. **b**. Top-5 fittest (highest mean AUPR) MRI classifier parameter solutions presented in descending fitness order. Top fitness (mean AUPR) values are presented in bold font.

### Custom Loss Function Terms Contribute to Classifier Performance

We sought to evaluate the relative contributions of the three-loss terms in our custom function. This was done to ensure that (1) each term contributes positively to model performance, and (2) that each parameter is well-optimized. In each of the three test scenarios, one loss term was omitted (two remained) and model AUPR was recorded (Figure 2a). In each case, the full loss term (i.e. that with all 3 terms) was top-performing. Next, statistical densities were visualized for each tradeoff parameter term versus fitness for all solutions evaluated by the GA (, ; Figure 2b). This plot reflects how focused the GA became on a certain parameter value as it evolved over generations. For example, for CT (Figure 2b top) we see that both and are honing in on values of approximately 1-10 and 500, respectively. Alternatively, in MRI (Figure 2b bottom) the parameter is more uniformly distributed across choices, potentially indicating a need for greater optimization. These relationships are also apparent in Figure 1a-b (e.g., has 1 unique solution and has 5 unique solutions for the top 5 networks). The vertical AUPR distributions seen in Figure 2b also reflect the fact that these three parameters work in concert to yield the highest performing loss functions.

**Figure 2.**
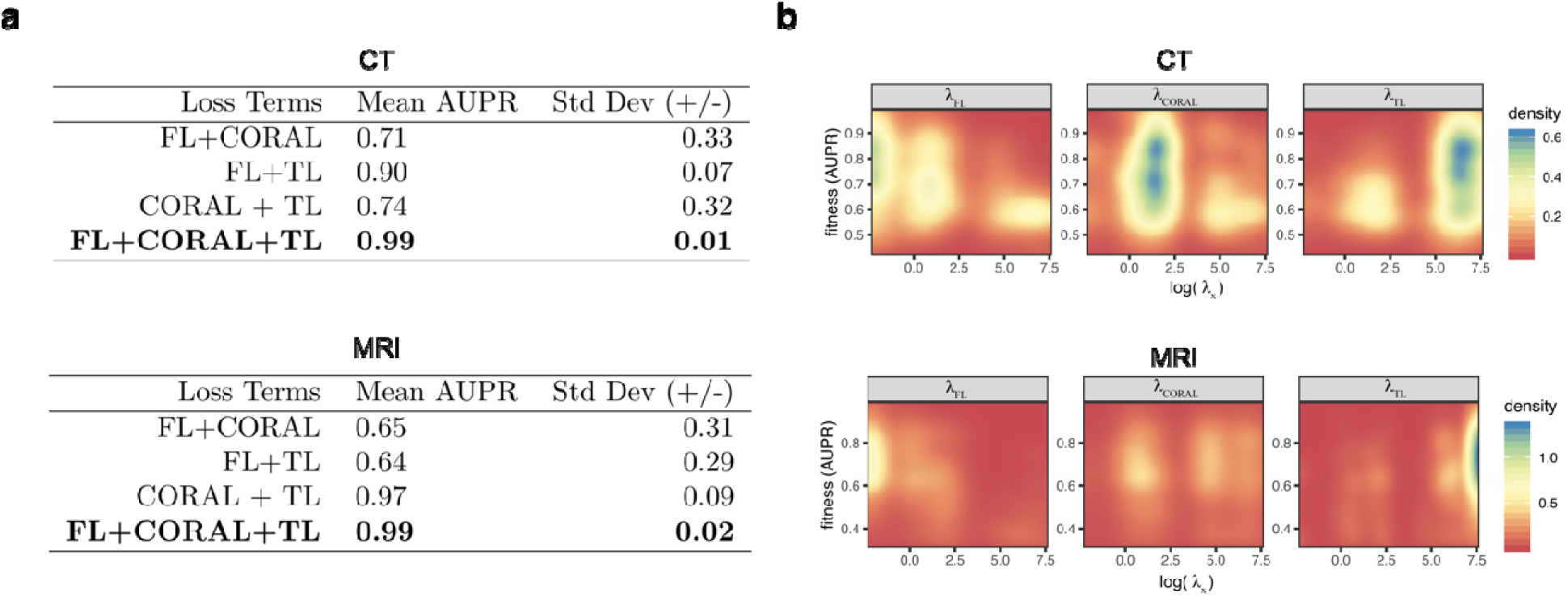
Loss Term Contributions to Classifier Performance. **a**. Optimized classifier performance with loss terms omitted for CT (top) and MRI (bottom) classifiers demonstrating the importance of the three loss terms. Highest performing scenarios presented in bold font. **b**. Two-dimensional statistical densities of log trade-off values versus mean AUPR (n=5) for (left to right),, parameters (CT: top; MRI: bottom). Blue pixels indicate greater density (i.e., frequency) while red pixels indicate lesser density.

## Discussion

This work demonstrates that advanced DL models can be utilized in small-scale contexts such as the diagnostic prediction of the brain tumor, Adamantinomatous Craniopharyngioma, from radiographic data. By concatenating the FL, CORAL, and TL loss terms and optimizing respective trade-off parameters via GA, we generated two DL classifiers with reproducible performance. Importantly, these classifiers significantly outperform published human expert performance (0.99 vs. 0.87) and do so by utilizing less than 80 examples. This stands in stark contrast to the previous DL work in medical fields, such a dermatology and diabetes which, due to their high incidence, have exemplar data numerous orders of magnitude larger in size (n>1×10^6^) than that which is available for pediatric brain tumors.

An intrinsic limitation to small-scale data analysis, which applies to this work, is that the training set itself is at risk of harboring sampling bias. Models trained on small datasets are therefore exposed to the risk of over-fitting and are therefore suspect with regard to reproducibility. To be clear, we must be hyper-critical of small data classifiers, especially in high-stakes scenarios, such as cancer diagnosis. To curtail the effect of this, we chose a novel loss function intended to be insensitive to outliers, and a robust fitness metric (AUPR) that we utilized via the 5-fold cross-validation approach, so that all data were inferred once. While the GA identified high-performing network solutions for both modalities, we maintain some reservations about model capacity due to the potential for artificial AUPR inflation from more separable groupings created by random chance.

Although we cannot perfectly estimate the novel inference capacity of our classifiers, these results demonstrate reproducible mean 5-fold cross-validation classification performance that is superior to published human expert accuracy, using both CT and MR images. We hypothesize that higher performance in the CT classification, relative to MRI, is due to the better sensitivity for microcalcifications on this modality. Such calcifications are a distinguishing characteristic of ACP relative to the other mass lesions in the differential diagnosis.^11^ Further, lower MRI performance may be due to the high degree of z-index variability (and therefore the quantity of information in each example) and could reflect a need to further optimize loss function parameters (also indicated in Figure 2b).

Potential next steps include further optimization of both modality classifiers, likely across different ranges of potential values based on the results in Figure 2b. Also, the parallel concatenation of both modality networks, thereby creating a scenario that trains on MRI and CT simultaneously, may yield a more robust classification framework. A further step forward would be to develop a fully interpretable classification tool, as an understanding of how a model fails is critical in high-stakes decision modeling. Lastly, the integration of our trained classifier into a lightweight web-based application for deployment to the broader medical community would enable easy integration into clinical workflows. As personalized medicine becomes a more attainable goal, the application of scalable frameworks and optimized DL models, such as those presented above, built on smaller and smaller datasets will be critical.

## Methods

### Computational Hardware and Software

All computational work was performed on a Graphics Processing Unit (GPU)-equipped High-Performance Intel CPU. Model construction, training, and evaluation were written and executed in Python 3.6, using the TensorFlow 2 (r2.1) machine learning framework obtained as the current (2020 March 1) “TF-nightly” docker environment, as recommended. Results were visualized using either the R tidyverse package or via Python’s Matplotlib module. ImageNet pre-trained ResNet V2-50 models were obtained through the TensorFlow Applications module.

### Image Acquisition

Deidentified radiographic data were obtained from consented patients via Children’s Hospital Colorado and all diagnoses were histologically determined by a board-certified neuropathologist via a standard protocol. Per United States Health and Human Services Regulation 45 CFR 46, this study was exempt from requiring Institutional Review Board approval. Preoperative non-contrast axial plane CT scans (N=28) and contrast-enhanced T1-weighted sagittal plane MRI scans (N=25) for subjects with Adamantinomatous Craniopharyngioma (ACP) were obtained in DICOM format. For the “NOTACP” control group, we collected the same modalities from patients with sellar/suprasellar masses with histologically confirmed diagnoses that are within the radiographic differential diagnosis for ACP (CT=47, MRI=24; Table 1). Genders were equally represented in both data classes (ACP: 27 females, 26 males; NOTACP: 24 females, 23 males).

**Table 1.** Statistical Frequencies of Diagnoses within NOTACP Class for CT and MRI Scenarios.

| Pathological Diagnosis | CT | MRI |
| --- | --- | --- |
|  | Frequency (n) | Frequency (n) |
| Arachnoid Cyst | 2 | 1 |
| Atypical Meningioma | 1 | – |
| Dermoid Cyst | 2 | – |
| Germinoma | 7 | 3 |
| Low-Grade Glioma | 2 | – |
| Lipoma | 1 | 1 |
| Mature Teratoma | 2 | 1 |
| Optic Glioma | 2 | 1 |
| Pilocytic Astrocytoma | 13 | 10 |
| Pilomyxoid Astrocytoma | 4 | 2 |
| Pituitary Adenoma | 3 | 2 |
| Prolactinoma | 4 | 2 |
| Renal Cell Carcinoma | 2 | 1 |
| Myelomeningocele (Shunted Hydrocephalus) | 1 | – |
| Subdural empyema | 1 | – |
| **Total (n)** | **47** | **24** |

### Image Preprocessing

For each DICOM series, pixel arrays were isolated using the pydicom python package. On a per slice basis, pixel arrays were standardized via mean removal and scaled to unit variance using the scikit-learn StandardScaler method. Next, for each slice, feature embeddings were generated by passing pixel arrays through the pre-trained ResNet V2-50 model. These embeddings were then saved as new files to be used downstream in the classifier, which decreased the computational memory and time required for model training.

### Network Architecture

In order to handle the varying dimensionality (i.e., channels, slices, z-index) within our dataset (*Z*_*CT*_ = 41±17, *Z*_*MRI*_ 51±52, Figure S1) in a manner that mirrors the processes used by human clinical experts, we utilized a sequence-based model (LSTM) in a bidirectional manner. The network is composed of two identical parallelized networks (source and target; Figure M1a), where the source network is trainable, the target network is not trainable (i.e., only source weights are updatable during training). Although the target network is capable of classifying target data (for early stopping purposes, as discussed in *Training & Evaluation* below), target network predictions are entirely disconnected from the objective loss function and therefore target prediction accuracy has no impact on training (Figure M1b).

**Figure M1.**
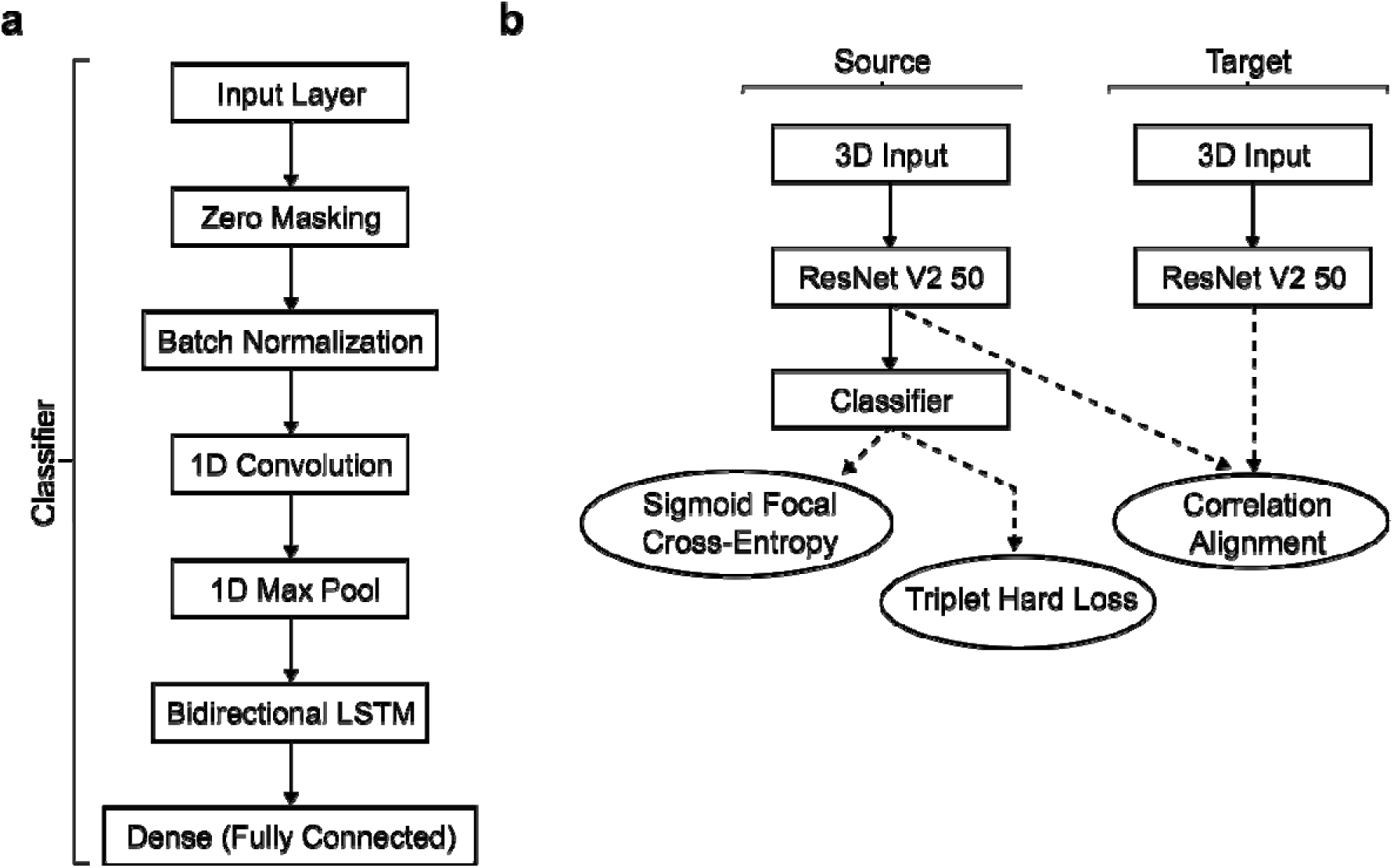
Network Architecture. **a**. Flow chart schematic for bidirectional LSTM network (classifier) utilized in this project. **b**. Parallelized source and target network flow chart with loss term connections (dashed lines).

### Custom Loss Function

To mitigate overfitting in model training, we utilized a custom loss function with 3 terms, each with a tradeoff parameter (a constant value used to weight loss terms;,,) that we optimized using a simple Genetic Algorithm (GA; Algorithm I, Methods). The first term used Sigmoid Focal Cross-Entropy (FL; implemented via TensorFlow AddOns) which was developed alongside the RetinaNet model.^8^ This loss function term is a modulated form of the standard cross-entropy loss function, defined as:

Where is standard cross-entropy, is the modulating term, and is the *tunable* focusing parameter. The FL term increases focus on samples that are not well-classified and de-emphasizes well-classified samples. It is therefore useful in class-imbalanced datasets.

The second loss term employed was the Triplet Hard Loss (TL; implemented via TensorFlow AddOns), which was first presented alongside the facial-recognition network, FaceNet.^7^ Intuitively, TL is an extension of the nearest neighbor (i.e., *k*-means) classification. This selects groups of three samples (triplets) where the first member, (anchor), is a given sample image, the second member is a positive-class instance,, and the last member is a negative-class instance,. TL is optimized by minimizing the distance between and, while maximizing the distance between. Mathematically this is represented as

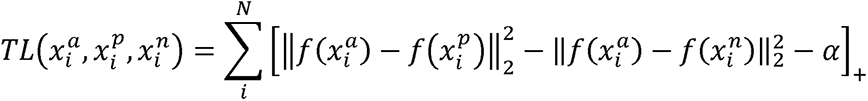

where *f*(*x*) ∈ ℝ^2^ is the data embedding in *d*-dimensional Euclidean space (constrained to the *d*-dimensional hypersphere; i.e., ‖ *f*(*x*) ‖_2_ = 1), and *α* is the margin enforced between positive and negative pairs. This loss term was added to our function due to the efficacy of discriminative feature learning of center-based loss functions.^6^

The final loss term was a slight modification of the domain adaptation method CORrelation ALignment (CORAL). Similar to the original CORAL method,^9^ our network utilized a pair of parallelized models (source and target). However, where the original CORAL method ties the entirety of these parallel models together and assesses the final fully-connected layer embeddings, we instead applied CORAL directly to the outputted feature embeddings (*X*_*s,embedding*_ and *X*_*t,embedding*_ for source and target feature embeddings, respectively) generated via ResNet V2-50. Therefore, our CORAL-like loss term is calculated as

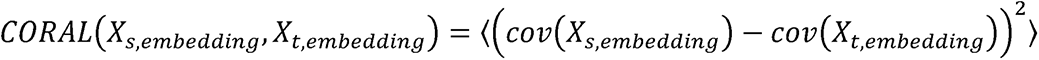

Where *cov*(·) is the standard covariance function. Combining these three terms together we get the final loss function to be optimized

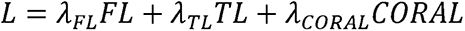

Where *λ*_*FL*_, *λ*_*TL*_, and *λ*_*CORAL*_ are trade-off parameters for the FL, TL, and CORAL loss terms to be optimized by genetic algorithm downstream.

### Genetic Algorithm Optimization

We sought to identify optimal batch size, initial learning rate, and tradeoff parameters to boost model performance. To achieve this, we utilized a simple Genetic Algorithm (GA) approach (Algorithm I). The GA iteratively selected from a pool of potential values for batch size, initial learning rate, and the tradeoff parameters. Fitness (5-fold mean Area Under Precision-Recall curve; AUPR) was monitored over generations to signal an early stop if no-improvement was seen for two generations (*n*_*gen,CT*_ = 4, *n*_*gen, MRI*_ = 6).

**Algorithm I.**
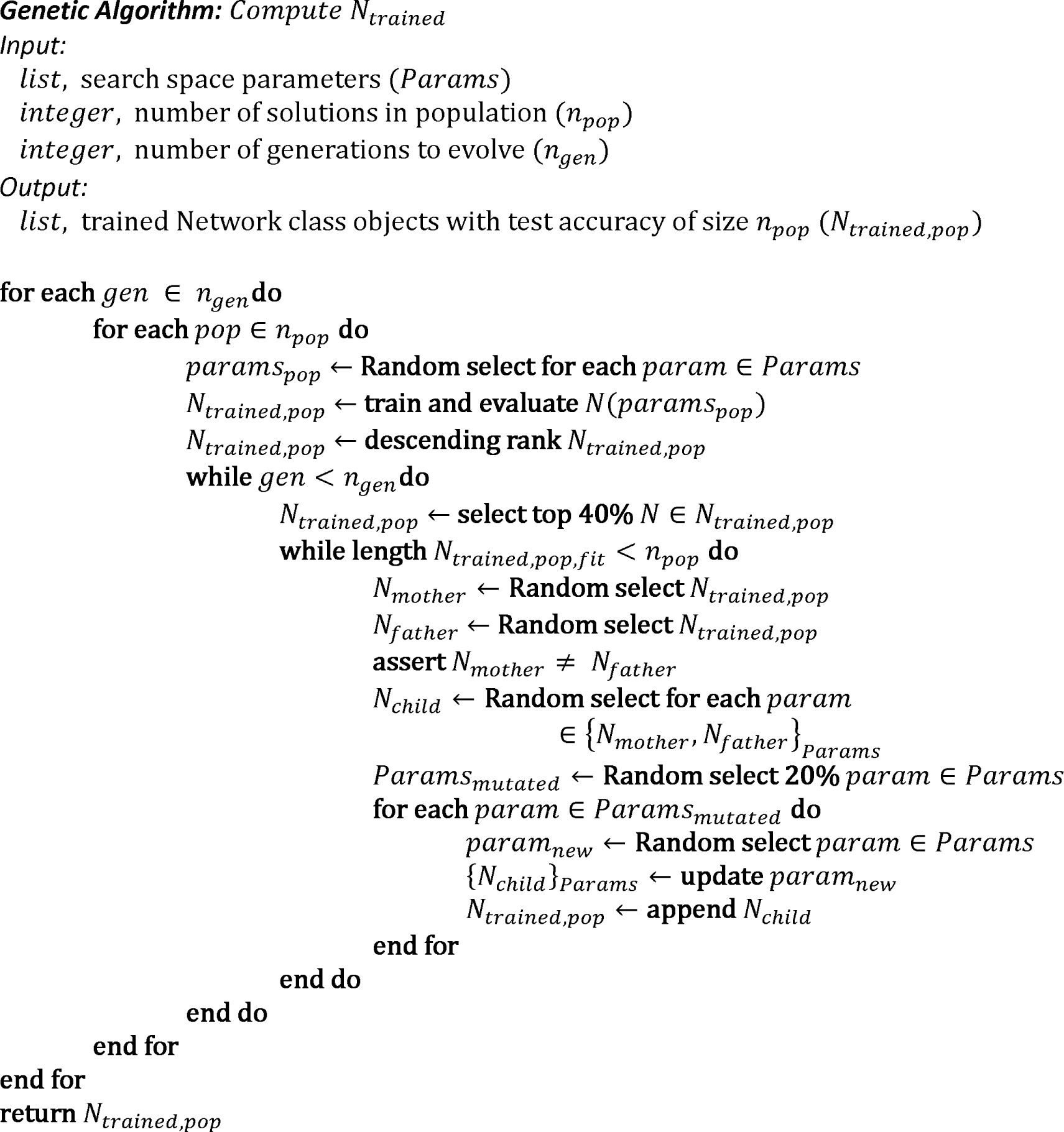

For batch size, we provided options of either N=4 or N=8. Although it is not uncommon to see much larger batch sizes in published work, we chose these two values due to the memory constraints of our HPC infrastructure. Furthermore, N=8 represents 10-16% of the cardinality for CT and MRI datasets, respectively. Increasing to N > 8 would likely lead to overfitting and decreased performance. The initial learning rate was set to a value within the set [10^−4^, 10^−5^, 10^−6^, 10^−7^]. These small learning rate values were chosen based on the premise that this step was fine-tuning the network, which was already leveraging ResNet V2-50 feature embeddings, and therefore required only small changes to optimize the objective function. Finally, loss trade-off parameters were chosen within the set [0.1, 1, 2, 5, 10, 100, 250, 500, 1000, 2000]. This list encompasses a range from fractionating a loss term to highly weighting it. In total, the genetic algorithm was executed for 10 generations with a population size of 100 (*N*_*evaluated*_ ≅ 1,000) and the number of potential solutions was 8,000.

### Training & Evaluation

To determine the model (inferential) capacity, we chose to use the 5-Fold Cross-Validation (5FCV) approach. Since we have parallelized source and target models, both datasets must have the same number of entries. However, the 5FCV approach segments the source and target data in a 4:1 manner. Therefore, for each iteration, we randomly oversample the target dataset such that source and target cardinality is equal.

The model was compiled using the stochastic gradient descent optimization algorithm implemented with a GA-determined initial learning rate subject to exponential weight decay (rate=0.96, decay steps=200). Maximum training duration was set at 500 epochs, and early stopping was enforced for target data evaluations exceeding a mean 5FCV of 0.87 Area Under the Precision-Recall curve (AUPR; this value equates to human performance) to minimize required training time. AUPR was chosen as the performance metric due to its known efficacy in representing accuracy in situations that are class imbalanced.

## Data Availability

Data is available upon reasonable request by contacting the Corresponding Author

## Supplemental Figures

**Figure S1.**
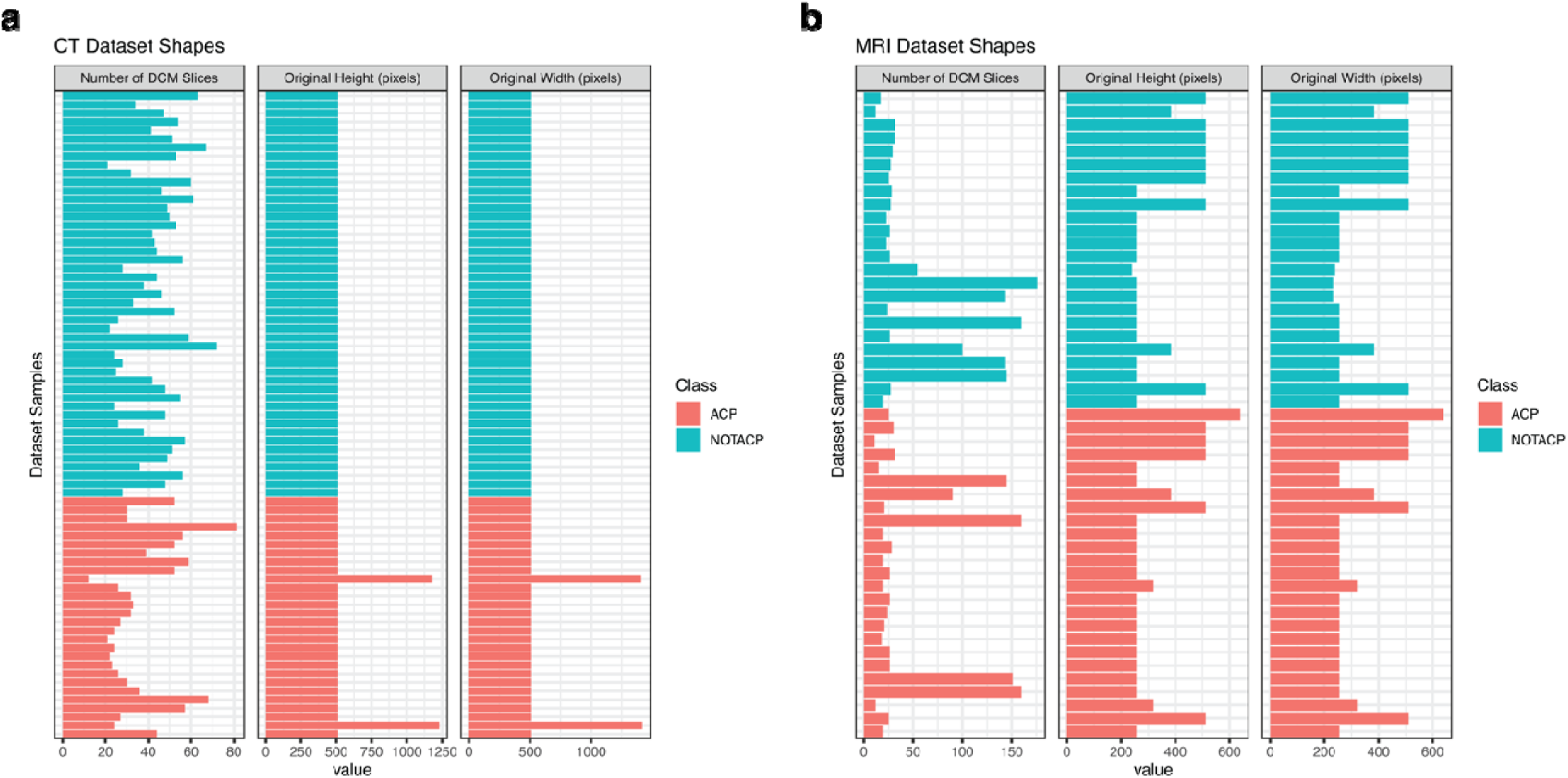
Radiographic dataset dimensionality for CT (**a**) and MRI (**b**) modalities.

## References

1. Russakovsky, O. et al. ImageNet Large Scale Visual Recognition Challenge. Int J Comput Vision 115, 211–252 (2015).

2. Gulshan, V. et al. Development and Validation of a Deep Learning Algorithm for Detection of Diabetic Retinopathy in Retinal Fundus Photographs. Jama (2016). doi:10.1001/jama.2016.17216

3. Srivastava, N., Hinton, G., Krizhevsky, A. & Sutskever, I. Dropout: A Simple Way to Prevent Neural Networks from Overfitting. 15, 1929–1958 (2014).

4. Lu, J. et al. Transfer learning using computational intelligence: A survey. Knowl-based Syst 80, 14–23 (2015).

5. Bengio, Y., Courville, A. & Vincent, P. Representation Learning: A Review and New Perspectives. (2012).

6. Wen, Y., Zhang, K., Li, Z., Qiao, Y. (2016). A Discriminative Feature Learning Approach for Deep Face Recognition

7. Schroff, F., Kalenichenko, D., Philbin, J. (2015). FaceNet: A Unified Embedding for Face Recognition and Clusteringhttps://dx.doi.org/10.1109/cvpr.2015.7298682

8. Lin, T., Goyal, P., Girshick, R., He, K., Dollár, P. (2017). Focal Loss for Dense Object Detectionhttps://arxiv.org/abs/1708.02002

9. Sun, B., Feng, J., Saenko, K. (2016). Correlation Alignment for Unsupervised Domain Adaptationhttps://arxiv.org/abs/1612.01939

10. Norris, G., Garcia, J., Hankinson, T., Handler, M., Foreman, N., Mirsky, D., Stence, N., Dorris, K., Green, A. (2019). Diagnostic accuracy of neuroimaging in pediatric optic chiasm/sellar/suprasellar tumors Pediatric Blood &Cancer https://dx.doi.org/10.1002/pbc.27680

11. Sartoretti-Schefer, S., Wichmann, W., Aguzzi, A., Valavanis, A. (1997). MR Differentiation of Adamantinomatous and Squamous-Papillary Craniopharyngiomas 18(), 77–87.

